# Differential Levels of Telomeric Oxidized Bases and TERRA Transcripts in Childhood Autism

**DOI:** 10.1101/2020.05.02.20088575

**Authors:** Mohammad Eftekhar, Yasin Panahi, Fahimeh Salasar Moghaddam, Mohammad Reza Eskandari, Hamid Pezeshk, Mehrdad Pedram

## Abstract

The underlying molecular mechanisms responsible for the etiology of autism and its sex-biased prevalence remain largely elusive. Abnormally shortened telomeres have recently been associated with autism. We have previously shown that children with non-syndromic autism exhibit a sexually dimorphic pattern of relative telomere length (RTL). Only male children with autism have significantly shorter RTLs than the healthy controls and paired siblings. Autistic females have substantially longer RTLs than autistic males. Aberrantly high levels of oxidative stress plays a fundamental role in the pathophysiology of autism, and telomeres are thought to be susceptible to oxidative damage due to their high guanine-repeat content. Employing a quantitative PCR (qPCR)-based method, telomeric oxidized base lesions were measured using genomic DNA extracted from saliva samples, and levels of telomeric RNA transcripts know as TERRA were evaluated using reverse transcriptase qPCR technique. Our data show that the autistic children exhibit substantially higher levels of oxidative base lesions at their telomeres than the healthy controls and paired siblings. Intriguingly, despite having significantly longer RTLs, female children with autism have even higher levels of telomeric oxidized bases than their male counterparts. Furthermore, despite having significantly shorter RTLs, the male children with autism exhibit lower levels of TERRA expression from the short arms of chromosomes 17 and X/P compared to their individually-matched healthy controls. These findings open a fresh angle into autism. Abnormal TL and high levels of telomeric oxidized bases may serve as biomarkers for childhood autism.

## Introduction

Childhood autism, a severe subgroup of autism spectrum disorders (ASD), is a lifelong pervasive neurodevelopmental disorder with an early onset characterized by deficits in reciprocal social interaction, impaired verbal and nonverbal communication, and restricted and repetitive behavior [1]. The number of boys diagnosed with autism is 4-5 times greater than that of girls [2]. Genetic factors play a major role in the etiology of autism, but the number of genomic variations characterized is overwhelmingly high and extremely heterogeneous with a high degree of variance in phenotypic presentations among different cases [3, 4]. Autism has a complex pathophysiology with a number of outstanding features and key roles for abnormal alterations in oxidative stress, immune and neuroimmune regulation and inflammation, metabolism, and mitochondrial function [5–7]. Despite heightened worldwide attention and highly significant advances in autism research in recent years, the cellular processes involved and the underlying molecular mechanisms responsible for the etiology and pathophysiology of autism remain largely elusive.

Two recent studies reported a strong association between abnormally shortened telomeres and autism implicating a potential role for telomere biology in the pathogenesis of autism [8, 9]. Telomeres are specialized and dynamic nucleoprotein structures at the terminal ends of the mammalian chromosomes composed of tandem repeats of double-stranded (ds) 5'-TTAGGG-3' extending for several thousand base pairs (typically 4-20 kb in humans) and 150-200 nucleotides of the so-called G-rich single-stranded (ss) overhang at the very end. The G-rich overhang is looped back in a large lariat structure with D- and T-loops. Telomeric sequences are associated with specific protein factors and enzyme complexes, which are also involved in maintaining telomere length (TL) and/or structure [10]. TL is an important factor in maintaining telomere structure and functions, which include protecting the chromosome ends as well as genome integrity [11]. Moreover, the specific factors associated with the telomeric sequences have a number of extra-telomeric functions [12]. Telomere shortening elevates dispersion of the telomere associated factors to chromosome internal loci [13], and critically short telomeres are a prelude to cellular senescence and apoptosis [14]. On the other hand, abnormally long telomeres are also indicative of unusual telomere structure and alternative telomere maintenance and function [10, 15].

Despite their classic characterization as constitutive heterochromatin and high levels of protection, telomeres are both susceptible to DNA damage and accessible to transcriptional machinery [13, 16, 17]. In addition to telomere erosion in replicating somatic cells, which lack the reverse transcriptase activity of telomerase, TL is dependent on a number of genetic plus internal and external environmental factors [18]. TL is impacted by oxidative stress induced damage and a variety of DNA-repair mechanisms under unique direction/influence of the protein factors and enzymes involved in telomere protection and maintenance [19, 20]. If not properly structured and/or protected, telomeres are particularly susceptible to oxidative damage because of their repetitive sequence and high consecutive guanine base content. In fact, 8-oxo-7,8-dihydrodeoxyguanosine (8-oxoG) is a major product of oxidative stress induced DNA damage [16, 21, 22]. Surprisingly, despite having a highly specialized heterochromatin structure and being devoid of classically-defined segments as genes, telomeres are transcribed into long non-coding RNAs referred to as TERRA [23]. TERRA molecules not only seem to be involved in telomere structure and TL regulation, but also capable of impacting the expression of sub-telomeric genes and a number of chromosome internal target genes, as well as a large network of interacting proteins [24]. TERRA expression levels depend on telomere length and structure [25, 26].

In line with earlier reports indicating key roles for oxidative stress and aberrant alterations in cellular redox status (*i.e*., anti-oxidant defense capacity) in autism [27–30], a comprehensive study from S. Jill James group provided convincing evidence on the elevated levels of both oxidized protein derivatives and genome-wide oxidized DNA adducts (including 8-oxo-dG) in children with autism but not their healthy control or paired sibling groups [5]. These observations were further confirmed by follow-up works from the same research lab by post-mortem analysis of the brain tissues from individual with autism and also a mouse model of autism [31, 32].

In our earlier report, we have presented evidence showing a sexually dimorphic pattern of average RTL status among children with non-syndromic autism. While autistic boys followed a homogenous pattern of abnormally shortened telomeres, there was no evidence of unusual telomere shortening in girls. In fact, in a sharp contrast to the autistic male children, some of the female children with autism had substantially longer average RTL than their matched healthy controls [33]. However, many aspects of telomere biology such as TL distribution among different chromosome ends, structure, transcriptional activity, and levels of telomeric oxidation remain unexplored. Here, by employing a quantitative PCR (qPCR)-based method previously described by Nathan O’Callaghan *et al*. [34], we examine the levels of telomeric oxidized base lesions among male and female autistic children. We also take a look at the levels of TERRA transcripts coming from certain chromosome ends that are of interest in autism. To our knowledge, this is the first report on the status of telomeric oxidation and TERRA transcription in ASD research.

## Results

### Subjects and Study Groups

Demographics and characteristics of the subjects including 24 children (14M:10F) with non-syndromic typical autism from independent simplex families, 10 apparently healthy paired siblings (8M:2F), and 24 sex-, age-, and location-matched healthy control children have been described in detail in our previous paper [33]. Clinically, all 14 male and 10 female autistic children were in “moderate to severe” category. Five out of 14 male children with autism and six out of 10 autistic female children were nonverbal. Eighteen (14M:4F) of the children with autism were drug naïve (i.e., first time diagnosis) at the time of sampling and four of the females had extended wash periods [33].

### The levels of oxidized residues in telomeric regions of autistic children are significantly higher than the healthy controls

Comparison of the average numbers of telomeric FPG-sensitive sites (i.e., oxidized base lesions) between different groups, after adjustment for covariates, showed that children with autism had significantly higher levels of telomeric base oxidation than both the healthy controls (β = 0.495, *p* = 0.0002) and apparently healthy paired siblings (β = 0.410, *p* = 0.003) (Fig. 1A, left). In a gender stratified manner, both male and female autistic children showed substantially higher levels of telomeric oxidized base lesions compared to their sex-, age-, and location-matched healthy controls (β = 0.506, *p* = 0.008; and β = 0.598, *p* = 0.002, respectively) (Fig. 1A, middle and right). Both male and female groups of children with autism also had significantly higher concentrations of telomeric oxidation compared with their respective paired siblings (β = 0.375, *p* = 0.048; and β = 0.622, *p* = 0.009, respectively). However, the difference for the female autistic children may not be very reliable because of the small number of paired siblings. Interestingly, the female autistic children had considerably higher levels of telomeric oxidation compared to their male counterparts, although this difference was of marginal statistical significance (β = 0.424, *p* = 0.053). It is also interesting to note that taking the sibling group out of the analysis further signifies the differences in the levels of telomeric oxidized bases between the children with autism either in combination or as male or female groups compared with their respective healthy control groups (β = 0.583, *p* = 0.00001; β = 0.611, *p* = 0.001; and β = 0.764, *p* = 0.001, respectively).

**Figure 1.**
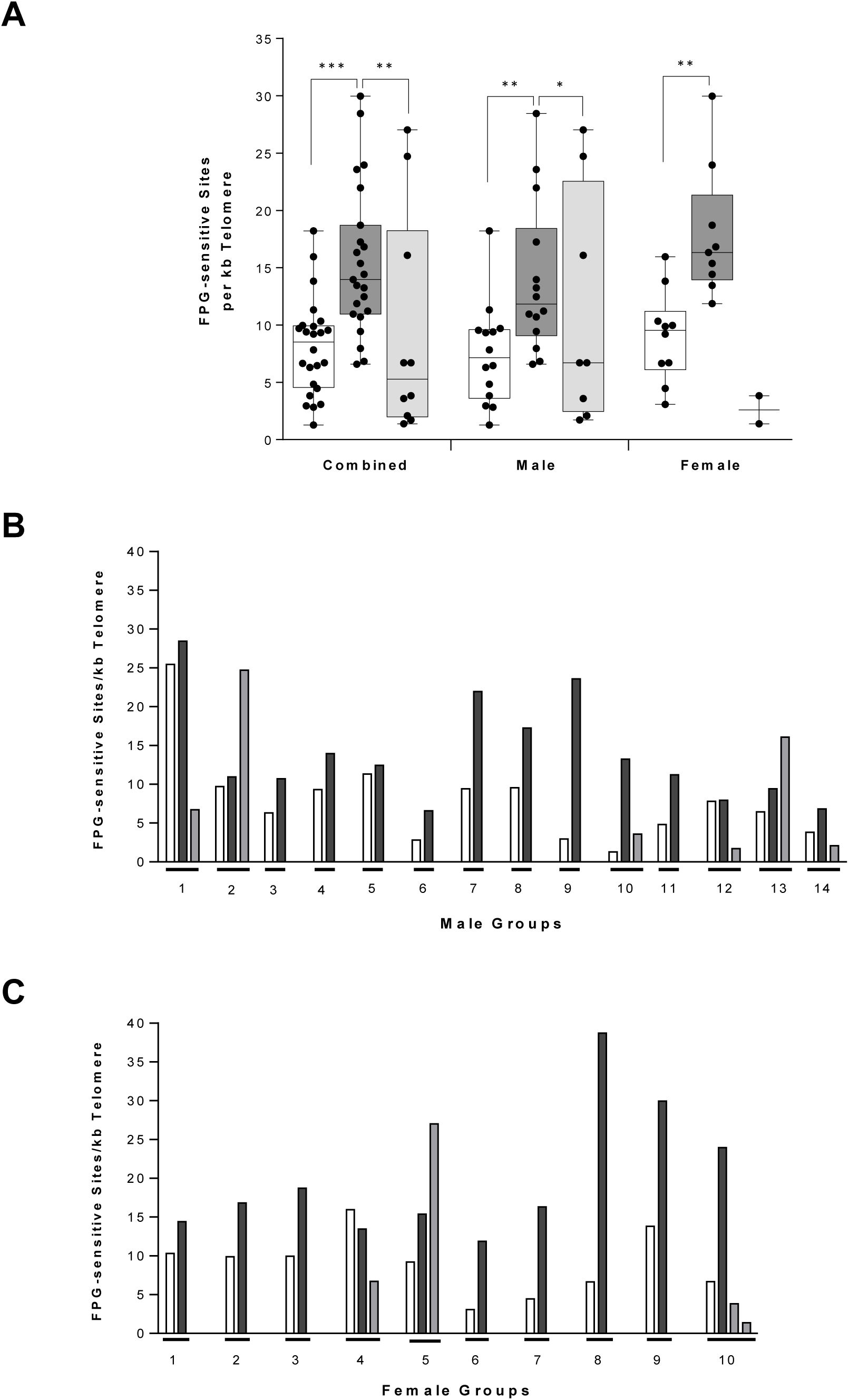
Concentration levels of telomeric oxidized bases in different clinical groups and individually matched subjects. Autistic subjects (in the middle), healthy siblings (to the right), and close neighboring healthy controls (to the left) are shown with dark, gray, and white boxes, respectively. **(A)** Comparison of the levels of oxidized bases per 1 kb of telomere sequences in children with idiopathic autism, their apparently healthy siblings, and healthy neighboring controls in both sexes (combined), and in male and female subjects exclusively. The bottom and top of each box represent the 25th and 75th percentiles, respectively, and the line bisecting each box shows the median value. Comparison of the telomeric oxidized base levels for patients, siblings, and healthy neighboring controls in male **(B)** and female **(C)** individually matched groups. Note: Siblings in the female groups 4 and 5 are male subjects. *, *p* < 0.05; *\*\*,p* < 0.01; *\*\*\*,p* < 0.001.

### Similar paterns of elevated levels of telomeric oxidized bases among autistic male and female children groups

Organizing the subjects as individually-matched groups, showed that with the exception of a very few cases, and in a sharp contrast to the sexually dimorphic patterns of average relative telomere lengths (RTLs) [33], both male and female autistic children exhibited a similar pattern of elevated telomeric oxidized bases compared with their individually-matched healthy controls and apparently healthy paired siblings (Fig. 1B and 1C). Also in contrast to the increased telomere attrition with age [33], the levels of telomeric oxidized base lesions remained rather constant in both autistic and control subjects with increasing age (Fig. 2). Similar to our observation on a lack of correlation between ADI-R total scores and RTL [33], there were no significant associations between the levels of telomeric FPG-sensitive sites and total ADI-R scores among the patients (Supplementary Material, Fig. S2) except for seemingly interesting contrasting trends for male vs. female nonverbal autistic children (Fig. S2, panel E).

**Figure 2.**
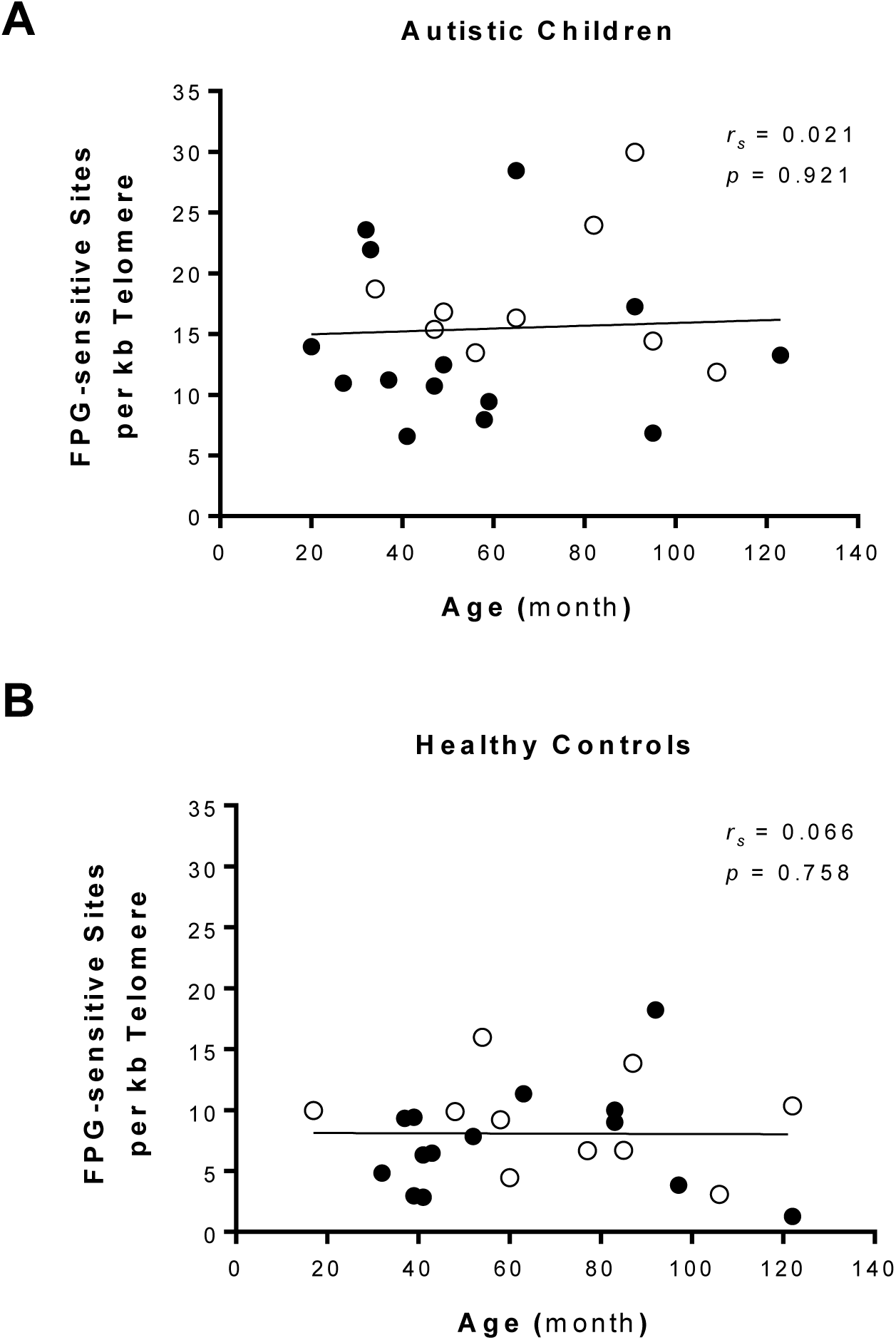
Telomeric oxidized base levels remain rather constant with age. Correlation between telomeric oxidized base levels and age in autistic children **(A)** and healthy controls **(B)** are shown. Black and open circles represent male and female individuals, respectively. Spearman’s correlation coefficient (*r_s_*) values and *P*-values are intercalated in each diagram area.

### Differential TERRA expression in male children with autism

We have previously reported a detailed evaluation of *GAPDH* and *YWHAZ* as appropriate combined reference genes for normalization of RT-qPCR data for saliva transcript level analysis in autistic male children [35]. We performed a similar systematic two-phased analysis among our autistic and contorl female samples. Although *GAPDH* and *YWHAZ* again were ranked as the best candidate reference genes among the panel of nine housekeeping genes examined, they did not pass the evaluation criteria. In fact, even a combination of 4 genes in the 2^nd^ phase of analysis did not seem good enough for RT-qPCR data normalization (Supplementray Material, Fig. S3). Thus, we limited our investigation of telomeric RNA transcription activity to the male samples alone.

We initially attempted to measure total TERRA levels in our male children groups following a protocol originally reported by Holzmann and colleagues [36]. However, despite careful treatment of the RNA samples with DNase I (see Materials and Methods), this approach did not produce a reliable outcome due to low levels of TERRA expression and background signals coming from even very small amounts of residual genomic DNA in the no-reverse transcription (NRT) control reactions (data not shown). Surprisingly, Holzmann and colleagues had not used any NRT controls for analysis of total TERRA expression in their study. Therefore, we switched our focus to measure TERRA molecules transcribed from specific chromosome ends 17p and XpYp, which are of interest in autism [37].

As shown in Figure 3 (panels A and D), both 17p- and XpYp-TERRA expression levels showed noticeable down-regulation in male children with autism compared with the healthy controls (4.8- and 29.6- fold, respectively). The expression levels of 17p- and XpYp-TERRA in individually-matched male groups are shown in Figures 3B and 3E. It should be noted that individual TERRA expression levels do not follow the pattern for the average RTLs shown in the accompanying paper [33]. However, the male children with autism, showed a significant decrease in both 17p- and XpYp-TERRA expression levels when compared with their individually matched healthy control samples [t(7) = −3.59, *p* = 0.009; and t(4) = −3.97, *p* = 0.016, respectively].

**Figure 3.**
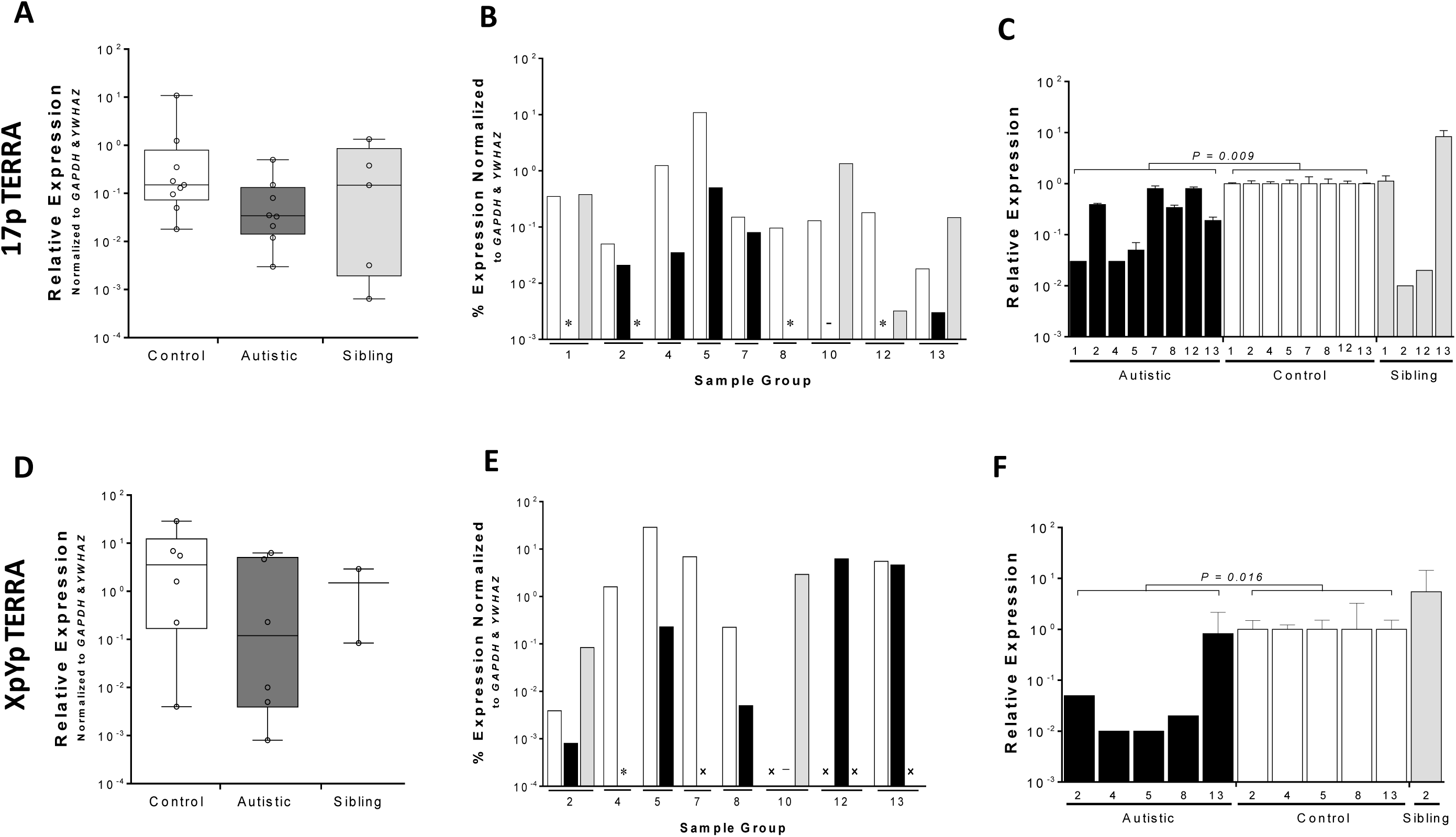
Expression levels of 17p- and XpYp-TERRA molecules in male subjects. Black bars are indicative of the autistic subjects. Healthy siblings and healthy neighboring controls are indicated with gray and white bars, respectively. The Box plots show the levels of 17p-TERRA **(A)** and XpYp-TERRA **(D)** molecules in male children with autism in comparison with the healthy controls and siblings. Data in panels A and D are presented as the median value and interquartile ranges, with the vertical bar representing the total range. Percent expression levels of 17p TERRA **(B)** and XpYp TERRA **(E)** in each sample, relative to the reference genes (*GAPDH* and *YWHAZ*), are shown for individual male subjects. The fold change of 17p TERRA **(C)** and XpYp TERRA **(F)** in each patient and his sibling relative to the patient’s respective healthy control is represented using one sample t-test. Signs: *, No signal detected for TERRA despite the *Cq* values both reference genes being reliable; -, Variable and/or unreliable signals; x, TERRA readings were considered as not reliable because of significant NRT signals (*i.e*., residual corresponding gDNA with average *Cq* value differences less than 4).

## Discussion

To our knowledge, this is the first evaluation of telomeric oxidation and transcription levels in children with autism and ASD in general. In our earlier research work, we showed that children with non-syndromic autism exhibited sexually dimorphic patterns of average RTL: while the male children with autism followed a homogeneous pattern of abnormally shortened telomeres, the autistic girls showed no sign of abnormal RTL shortening. Moreover, some female autistic cases had substantially longer RTLs than the healthy controls [33]. Here, we show that the levels of telomeric oxidized base lesions are substantially higher in autistic children than the paired siblings and healthy controls. Despite having longer telomeres, female autistic children show even higher levels of telomeric oxidation levels than their male counterparts. Also somewhat surprising, despite having shorter RTLs, autistic male children show a significant decrease in 17p- and XpYp-TERRA expression levels when compared with their individually-matched healthy controls. We will discuss some possible implications of these seemingly paradoxical observations below.

Because of their repetitive sequence and strings of GGG, telomeres are considered as “hot zones” for oxidative damage [16, 38, 39]. As outlined above, our data show substantially higher levels of telomeric oxidized base lesions in both male and female children with autism. Thus, based on the conventional view on the association of increase in oxidative stress with accelerated telomere shortening, one would expect that autistic children to exhibit abnormally shortened telomeres. While this prediction fits well with our observations for the autistic male children, it sharply contrasts with our data from the autistic females. Previously, we provided a detailed argument explaining how differences in sex biology and hormones could lay the foundations for sexual dimorphism in RTL between male and female children with autism [33]. How come then autistic girls with longer RTL have even higher levels of telomeric oxidized base lesions than their male counterparts? First, we would like to remind the reader that we do not know of the actual length distribution of different chromosome-specific telomeres. It is therefore possible that the longer average RTL in autistic girls to be the result of certain abnormally long chromosome ends not all telomeres. Second, we do not know the exact composition of the oxidized bases within the telomeres, which are composed of 25% G and 25% T for most of their length as dsDNA, followed by 50% G and 33.3% T within the small but crucial ssDNA overhang at the very end. It is true that the bacterial enzyme FPG has a preference for oxidized purines, in particular 8-oxoG and its derivatives, but it also acts on thymine glycol (Tg) and has apurinic/apyrimidinic lyase activity, introducing ssDNA breaks [40]. In other words, FPG-sensitive sites could easily contain oxidized bases other than 8-oxoG. Most notably, it has been shown that Tg residues, which are a common oxidative product of thymine [21, 41] have significant impacts on telomere length, stability, and maintenance [42–44]. Last but not least, we do not know the exact locations and origins of telomeric oxidized bases. Nor do we know the structures of the autistic telomeres and the type of protein factors interacting with the oxidized bases. The nature, distribution, and impact of different oxidized lesions at human telomeres remain largely elusive and poorly understood. Different combinations of oxidized bases and lesions at telomeres, interactions with the shelterin proteins and various DNA repair intermediates could result in contrasting telomere length and integrity outcomes [19, 20].

The most studied oxidized base 8-oxoG itself is prone to further oxidation and turning into a number of hydantoin derivatives [45]. It is also important to note, however, that even if we assume that 8-oxoG and its derivatives make up the majority of oxidized lesions at autistic telomeres, it is difficult to predict the outcome on telomere length and structure without knowing the composition and locations of these residues, their sources, the composition of other oxidized lesions, and interacting factors and chromatin context [46]. Telomeres are highly unique chromatin entities capable of forming alternate structures including T- and D-loops and G-quadruplexes (G4), interacting with specially coded histones and histone variants, sheltering proteins, and chromatin remodeling complexes. The impact and processing of the 8-oxoG and its derivatives and other oxidized lesions appear to be distinctly different at the telomeres depending on their origins, compositions, and contexts [19, 20]. Moreover, although 8-oxoG is conventionally regarded as a moderate mutagen and a biomarker of cellular oxidative stress, recent reports suggest that it functions as an “epigenetic-like” mark at chromosome internal sites, and its impact on chromatin structure and transcription is very much dependent on the genomic context within which it is situated and the interacting protein factors involved [46]. Of particular interest is the evidence showing 8-oxoG residues occurring at specific G-rich regulatory elements and their ability to increase transcription by promoter G4 formation mediated by the base excision repair machinery [46, 47]. Recent studies by Opresko, Myong, and colleagues have revealed some interesting insights into the structural impact and regulatory mechanisms of the two most common oxidized bases, 8-oxoG and Tg, within the telomeric ssDNA 3’-overhang on human telomerase activity and telomere elongation using tailored ssDNA synthesized oligonucleotides in human HEK-293 and HeLa cells [44, 48]. Interestingly, while insertion of the oxidized dGTP free nucleotides by telomerase inhibits its reverse transcriptase activity and leads to telomere shortening, the 8-oxoG residues that already exist in the 3’-overhang sequence disturb the G4 structures and simulate telomerase loading and activity leading to telomere elongation [48]. Also interestingly, in contrast to the *in vitro* data showing Tg acting as a strong block to DNA replication [49] and evidence suggesting that Tg residues cause telomere fragility in transgenic mice lacking the NTH1 glycosylase, the enzyme which removes Tg from telomeric sequences [43], Myong and colleagues have shown that Tg lesions in the G-rich telomeric ssDNA oligonucleotide actually stimulate telomerase binding and telomere elongation [44]. The status, compositions, and functions of oxidized lesions at the native mammalian telomeres under physiological conditions are simply not known.

A study by Jill James’s group has shown significantly higher levels of genomewide oxidative markers (including 8-oxo-dG) in the autism cerebellum and Brodmann area 22 [31], which is part of the superior temporal gyrus in the left hemisphere and plays a crucial role in speech processing [50]. We see no evidence of higher levels of telomeric oxidized bases among our nonverbal group of patients. Interestingly, Rose *et al*. also concluded that since they saw no correlation between the levels of oxidative biomarkers and age, this was a chronic condition in autism [31]. We, too, found no association between age and the level of telomeric oxidized bases among either the children with autism (Fig. 2B) or the healthy controls (Fig. 2A). Taken together, this brings up the possibility that the oxidative status of telomeric regions may have been set during early/critical period of neurodevelopment. Moreover, based on our data showing an inverse correlation between age and RTL [33], it can be concluded that the overall distribution of oxidized bases remains about even throughout most of the telomeric dsDNA.

Another seemingly paradoxical observation among our findings is that despite having significantly shorter average RTL in male children with autism (M.Pedram and colleagues, manuscript in preparation), they had lower levels of 17p- and XpYp-TERRA expression, which may look contrary to the expected inverse correlation between telomere length and TERRA expression [25]. However, in addition to not knowing the telomere length status at the short arms of chromosomes 17, X, and Y in our samples, we are also unaware of the structure and epigenetic status of these telomeres. It is possible that despite having shorter than normal average telomere length, the male children with autism have longer than usual telomeres at certain chromosome ends. Furthermore, having shorter TL does not necessarily translate into elevated levels of TERRA transcription. A very interesting and real-life example of this is demonstrated in Cockayne syndrome, a rare hereditary human disorder, marked by severe segmental premature aging and progressive neurological degeneration. Most Cockayne syndrome patients carry a deleterious mutation in the Cockayne syndrome group B (CSB) gene that encodes for a large nuclear protein implicated in base excision repair, transcription regulation, and chromatin remodeling [51]. It has been shown that CSB protein localizes to only a small subset of telomeres, and that CSB loss/deficit leads to telomere shortening as well as lower levels of TERRA transcription [52]. Another interesting case supporting non-random TL distribution and maintenance among individual chromosome ends has been demonstrated in patients with ICF (immunodeficiency, centromeric instability and facial anomalies) syndrome, type I [53]. It is also interesting to note that a recent study on mouse embryonic stem cells has revealed that in addition to binding to telomeres and stabilizing them in *cis*, TERRA molecules bind to a number of select genes and also interact with a sizable proteome including a number of very important telomeric proteins, epigenetic factors, and non-telomeric enzymes such as the RNA helicase ATRX and chromatin modifier CHD8 [24]. The gene encoding CHD8 is frequently mutated in ASD, and in fact, CHD8 appears to be a master regulator of ASD risk genes [4]. Interestingly, a recent study shows that a human *CHD8* mutation leads to sexually dimorphic behavioral phenotypes (with male-preponderant deficits) and neuronal activity in transgenic mice [54].

Our observations on the sexually dimorphic patterns of telomere length in children with non-syndromic autism [33], as well as differential 17p- and XpYp-TERRA expression levels in autistic male children, could provide important insights for deciphering telomere dynamics and the underlying molecular mechanisms of childhood autism. So many interesting and important questions come to surface: What is the status of TL at specific chromosome ends in male vs. female children with autism? What is the composition of telomeric oxidized bases in male vs. female autistic children vs. that of healthy controls? What factors interact with these oxidized bases/telomeres? What are the actual impacts of 8-oxoG and Tg at native telomeres in autistic children vs. healthy controls? What is the status of DNA methylation in subtelomeric regions of male vs. female children with autism vs. that of healthy controls? What is the implication of differential chromosome-specific TERRA expressions in male children with autism? What is the status of specific TERRA expression levels in female children with autism? What would be the impact of differential TERRA expression in autistic male (and perhaps, also female) children vs. controls on telomere structure, subtelomeric gene expression, and various TERRA target genes and interacting proteins in trans (for example, the chromatin modifier CHD8)? Do TERRA molecules play a part in explaining sexual dimorphism and male-bias in autism? Answering these questions could provide important and highly valuable insights and clues for deciphering the underlying molecular mechanisms involved in the etiology of autism and also provide a better understanding of the role of telomere biology in the molecular mechanisms responsible for sex bias and shared pathophysiology in autism.

## Materials and Methods

### Subjects and Sampling Procedure

This work follows our previous study on children with non-syndromic autism including: 24 children (14 male and 10 female) with non-syndromic autism (61.50 ± 27.65 mo), 24 sex-, age-, and location-matched healthy control children (66.08 ± 28.08 mo) as well as 10 apparently healthy siblings (eight males and two females, 80.40 ± 55.44 mo) [33]. Briefly, children with autism were diagnosed by two qualified clinicians, a child neurologist and a specialized psychiatrist, and the diagnosis of autism was further confirmed by Autism Diagnostic Interview-Revised (ADI-R) [55]. The Majority (18 subjects) of the children with autism (including all of the males) were antipsychotic drug naïve at the time of diagnosis and saliva sampling and six of the autistic females with treatment history had extended wash out periods prior to sampling. The present study was approved by Zanjan University of Medical Sciences (ZUMS) Research Ethics Committee (ZUMS.REC.1392.97 and REC.1394.265), it was conducted in accordance with the Declaration of Helsinki, and all children had signed informed consent provided by their parents or caregivers.

Saliva sampling was done using the Oragene collection kits (DNA genoTeK Inc., Ottawa, Ontario, Canada) as described previously [35]. OG-250 saliva-collection kits were used for genomic DNA (gDNA) extraction, and RE-100 kits were used for RNA extraction. Collected saliva samples from different cities and provinces were kept at room temperature for no longer than a week and were subsequently transferred to the central laboratory at ZUMS for appropriate processing.

### Telomeric 8-oxoG/Oxidized Base Measurement

The concentration of oxidized base lesions in telomeric regions was evaluated using a qPCR-based technique introduced by Nathan O’Callaghan and colleagues [34]. This method combines gDNA digestion with the formamidopyrimidine (FPG) enzyme [40], a bacterial DNA glycosylase that is involved in DNA repair through Base Excision Repair (BER) mechanism [56], and a variation of the qPCR telomere measurement technique originally devised by Richard Cawthon [57]. FPG preferably cuts the oxidized purines, in particular 8-oxoG [58]. The difference in PCR threshold cycles between DNA templates completely digested by FPG and undigested DNA is then calculated (*i.e*., Δ*C_q_* = *C_q_*digested - *C_q_*undigested).

### 8-oxoG Standard Curve Preparation

Four 84-mer oligomers of 14 5'-TTAGGG-3' repeats (as forward oligomers) containing either zero, one, two, or four 8-oxoG bases each and also a reverse oligomer (an 84-mer sequence of 14 5'- CCCTAA-3' repeats) [34] (Supplementary Material, Table S1), were purchased from Jena Bioscience (Jena, Thuringia, Germany). To prepare duplex standard oligonucleotides, each of the forward oligomers were mixed with the reverse oligomer in an equal molar ratio separately, followed by incubation at 95°C for 10 min and then cooling at room temperature for 30 min. The enzymatic digestions were performed using FPG (New England BioLabs, Hitchin, UK) on each of the ds-oligonucleotides containing zero, one, two and four 8-oxoG bases (Tel 0, Tel 1, Tel 2, and Tel 4, respectively) in microtubes. 200 ng of each duplex standard oligomers were incubated with 8 units of FPG in 1X NEBuffer 1 and 1X BSA. In case of undigested oligomers as controls, FPG was replaced with ddH2O in the reaction tubes. The reactions were set up on ice, and all microtubes/reactions (digested and undigested) were then incubated at 37°C for 4.5 hr, followed by an enzyme inactivation step at 60°C for 10 min. To evaluate complete digestion, a small portion of each sample was run on a 2.5% agarose gel.

Following enzymatic digestion with FPG (New England BioLabs, Hitchin, UK), amplification of the oligomers was performed using ABI Step One Plus Real-Time PCR instrument (Applied Biosystems, Foster City, CA, USA). Primer sequences were as follows: teloF: 5′- CGGTTTGTTTGGGTTTGGGTTTGGGTTTGGGTTTGGGTT-3′ and teloR: 5′- GGCTTGCCTTACCCTTACCCTTACCCTTACCCTTACCCT-3′ [59]. qPCR reactions were set up in a final volume of 20 µL containing 40 pg of digested or undigested standard oligomers, 1X SYBR Green master mix, high ROX (Ampliqon, Odense, Denmark), and 100 nM of each primer. PCR amplifications were performed in triplicates with an initial enzyme activation step at 95°C for 10 min, followed by 40 cycles of 95°C for 15 sec and 60°C for 30 sec. Values for ΔC_q_ (*C_q_* treated - *C_q_* untreated) were then calculated and used for preparing the 8-oxoG standard curve (Supplementary Material, Fig. S1).

### Measurement of Telomeric Oxidized Base Lesions in gDNA Samples

Genomic DNA (gDNA) was extracted from whole saliva samples, using the manufacturer’s protocol (OG-250; DNA genoTeK Inc., Ottawa, Ontario, Canada) with slight modification [33]. 240 ng of each gDNA sample was incubated with 12 units of FPG in 1X NEBuffer 1 and 1X BSA. To prepare undigested controls, the FPG enzyme replaced with an equivalent volume of ddH_2_O. Reactions were set up on ice and all microtubes/reactions (digested and undigested) were incubated at 37°C overnight followed by an enzyme inactivation step at 60°C for 10 min.

Amplification of telomeric regions in digested and undigested gDNA was performed as described in the previous section. PCR reactions were prepared in a final volume of 20 μL containing 20 ng of digested or undigested genomic DNA, 1X SYBR Green master mix, high ROX (Ampliqon, Odense, Denmark), 100 nM teloF, and 100 nM teloR primers. PCR amplifications were performed in triplicates with an initial enzyme activation step at 95°C for 10 min followed by 40 cycles of 95°C for 15 sec and 60°C for 30 sec. Values for Δ*C_q_* (*C_q_* treated - *C_q_* untreated) were then calculated. The number of oxidized base lesions (*i.e*., FPG-sensitive sites) per 1 kb of telomeric DNA were then determined using the 8-oxoG standard curve.

### Telomeric RNA (TERRA) Primer Set Selection

In an attempt to determine total (i.e., not chromosome-specific) TERRA levels, we used the telomere primer sets originally introduced by Richard Cawthon [60]. Chromosome-specific TERRA primer sequences for the chromosome ends 17p and XpYp were selected from previous publications [23, 61]: 17p TERRA-F, 5′-CCACAACCCCACCAGAAAGA-3′; 17p TERRA-R, 5′- GCGCGTCCGGAGTTTG-3′; XpYp TERRA-F, 5′ - GAATCCACGGATTGCTTTGTGTACTT-3′; and XpYp TERRA-R, 5′ - CCTCAGCCTCTCAACCTGCTTGG-3′.

### RNA Extraction and Analysis of TERRA Transcript Levels with RT-qPCR

Total RNA was extracted from whole saliva as described in detail previously [35]. Briefly, total RNA was reverse transcribed to Complementary DNA (cDNA) by using the PrimeScriptTM RT reagent Kit (TaKaRa Inc., Shiga, Japan) following the manufacturer’s instructions. To eliminate potential residual genomic DNA, each 500-ng RNA sample were treated with 0.5U RNase-free DNase I (1U/ µL) (Thermo Scientific, Waltham, MA, USA) prior to cDNA synthesis. TERRA transcripts originating from chromosome ends 17p and XpYp were amplified using an ABI Step One Plus Real-Time PCR System (Applied Biosystems, Foster City, CA, USA). Each 20-µL PCR reaction for either 17p or XpYp TERRA amplification was composed of 25 ng and 50 ng of cDNA, respectively, 10 µL of Power SYBR Green master mix (Applied Biosystems, Foster City, CA, USA), and 150 nM of each forward and reverse primers. PCR amplifications were performed in triplicates with an initial enzyme activation step at 95°C for 10 min, followed by: 50 cycles of 95°C for 15 s, either 6°C (for XpYp TERRA) or 62°C (for 17p TERRA) for 30 s, and 72°C for 1 min.

Selection and validation of *GAPDH* and *YWHAZ* combination as the most stable and suitable reference genes for analysis of the salivary transcriptome in male children with non-syndromic autism has been described previously [35]. A similar analysis of the cDNAs derived from female saliva samples showed that although *GAPDH* and *YWHAZ* were still the best among a panel of nine candidate reference genes examined, they would not be sufficient for proper normalization of the RT-qPCR data obtained from female subjects (Supplementary Material, Fig. S3). Therefore, the RT-qPCR data were normalized by using the geometric mean of GAPDH and YWHAZ Cq values in male samples only. All reactions were run in triplicates. No-template (NTC) and no-reverse transcriptase (NRT) controls were included in each run.

### Statistical Analysis

Distributions of telomeric oxidized base levels among different clinical groups were non-normal based on their respective histograms and/or Shapiro-Wilk tests. To achieve normal distribution, data points were first natural log transformed and then compared by using multiple linear regression. To compare the mean values of telomeric oxidized base levels among different clinical groups, groups were coded as dummy variable and incorporated into the regression analysis with adjustment for covariates including sample’s age, sex, maternal ages at birth, and birth weight. Spearman’s rho correlations were used to examine the association between telomeric oxidized base levels and ADI-R scores. To evaluate TERRA expression levels among different clinical groups, a One-sample t-test was used, and each affected subject was calibrated relative to his/her respective healthy neighboring control to minimize the effect of environment. To avoid skewness, relative expression level values were first transformed to log values to assure a normal distribution before performing the One-sample t-test.

All statistical tests were two-tailed, and the P-values below 0.05 (*p <* 0.05) were considered as statistically significant. Statistical analysis was performed using the SPSS software, ver. 22.0 (IBM Corp., Armonk, NY, U.S.A.). Scatter plots and bar graphs were made using GraphPad Prism, ver. 6.07 (GraphPad Software Inc. La Jolla, CA, U.S.A.).

## Data Availability

The only additional relevant data referred to in the manuscript could be obtained from our earlier manuscript submitted to medRxiv shown below:
Panahi, Y., Salasar Moghaddam, F., Babaei, K., Eftekhar, M., Shervin Badv, R., Eskandari, M.R., Vafaee-Shahi, M., Pezeshk, H. and Pedram, M. (2020) Sexual Dimorphism in Telomere Length in Childhood Autism. MedRxive, doi: 10.1101/2020.04.30.20074765.

## Supplementary Material

A composite PDF document containing the following:

One Table (Table S1); Three Figures (Figs. S1-S3); and References (total of 3).

## Abbreviations

8-oxoG: 8-oxo-dihydro-2'-deoxyguanosine
ASD: Autism spectrum disorders
ADI-R: Autism Diagnostic Interview-Revised
BER: Base Exision Repair
*Cq*: Quantification Cycle
ds: doublestranded
DSM-IV-TR: Diagnostic and Statistical Manual of Mental Disorders, Fourth Edition, Text Revision
FPG: Formamidopyridine DNA Glycosylase
*GAPDH*: Glyceraldehyde-3-phosphate dehydrogenase gene
G4: G-quadruplex
ICD-10: International Statistical Classification of Diseases and Related Health Problems version 10
NRT: No Reverse Transcriptase
NTC: No Template Control
ROS: Reactive Oxygen Species
RT: Reverse Transcriptase
RTL: Relative Telomere Length
RT-qPCR: RT Quantitative real-time PCR
ss: single-stranded
TERRA: Telomeric Repeat Containing RNA
Tg: Thymine glycol
TL: Telomere Length
*YWHAZ*: Tyrosine 3 monooxygenase activation, zeta plolypeptide gene

## Funding

This work was supported by Zanjan University of Medical Sciences (ZUMS) grant numbers A-12-534-1, A-12-534-9, and it had the approval of the ZUMS Research Ethics Committee (ZUMS.REC.1392.97 and REC.1394.265).

* Please, note that ZUMS is an academic institution. The present work/research is conducted independent of the Iranian government agencies.

## Acknowledgements

We would like to thank Reza Shervin Badv, M.D. (Dept. of Pediatric Neurology, School of Medicine, Tehran University of Medical Sciences), Mohammad Vafaee-Shahi, M.D. (Growth and Development Research Center, Institute of Endocrinology and Metabolism, Iran University of Medical Sciences) for the initial recruitment and clinical examination of some of the autistic children, and clinical follow-ups.

## References

1 Lai, M.C., Lombardo, M.V. and Baron-Cohen, S. (2014) Autism. Lancet, 383, 896–910.

2 Fombonne, E. (2009) Epidemiology of pervasive developmental disorders. Pediatr Res, 65, 591–598.

3 Tick, B., Bolton, P., Happe, F., Rutter, M. and Rijsdijk, F. (2016) Heritability of autism spectrum disorders: a meta-analysis of twin studies. J Child Psychol Psychiatry, 57, 585–595.

4 Arnett, A.B., Trinh, S. and Bernier, R.A. (2018) The state of research on the genetics of autism spectrum disorder: methodological, clinical and conceptual progress. Curr Opin Psychol, 27, 1–5.

5 Melnyk, S., Fuchs, G.J., Schulz, E., Lopez, M., Kahler, S.G., Fussell, J.J., Bellando, J., Pavliv, O., Rose, S., Seidel, L. et al. (2012) Metabolic imbalance associated with methylation dysregulation and oxidative damage in children with autism. J Autism Dev Disord, 42, 367–377.

6 Rossignol, D.A. and Frye, R.E. (2014) Evidence linking oxidative stress, mitochondrial dysfunction, and inflammation in the brain of individuals with autism. Front Physiol, 5, 150.

7 McCarthy, M.M. and Wright, C.L. (2017) Convergence of Sex Differences and the Neuroimmune System in Autism Spectrum Disorder. Biol Psychiatry, 81, 402–410.

8 Li, Z., Tang, J., Li, H., Chen, S., He, Y., Liao, Y., Wei, Z., Wan, G., Xiang, X., Xia, K. et al. (2014) Shorter telomere length in peripheral blood leukocytes is associated with childhood autism. Sci Rep, 4, 7073.

9 Nelson, C.A., Varcin, K.J., Coman, N.K., De Vivo, I. and Tager-Flusberg, H. (2015) Shortened Telomeres in Families With a Propensity to Autism. J Am Acad Child Adolesc Psychiatry, 54, 588–594.

10 Blasco, M.A. (2005) Telomeres and human disease: ageing, cancer and beyond. Nat Rev Genet, 6, 611–622.

11 O’Sullivan, R.J. and Karlseder, J. (2010) Telomeres: protecting chromosomes against genome instability. Nat Rev Mol Cell Biol, 11, 171–181.

12 Martinez, P. and Blasco, M.A. (2011) Telomeric and extra-telomeric roles for telomerase and the telomere-binding proteins. Nat Rev Cancer, 11, 161–176.

13 Ye, J., Renault, V.M., Jamet, K. and Gilson, E. (2014) Transcriptional outcome of telomere signalling. Nat Rev Genet, 15, 491–503.

14 Lechel, A., Satyanarayana, A., Ju, Z., Plentz, R.R., Schaetzlein, S., Rudolph, C., Wilkens, L., Wiemann, S.U., Saretzki, G., Malek, N.P. et al. (2005) The cellular level of telomere dysfunction determines induction of senescence or apoptosis in vivo. EMBO Rep, 6, 275–281.

15 Stanley, S.E. and Armanios, M. (2015) The short and long telomere syndromes: paired paradigms for molecular medicine. Curr Opin Genet Dev, 33, 1–9.

16 Oikawa, S. and Kawanishi, S. (1999) Site-specific DNA damage at GGG sequence by oxidative stress may accelerate telomere shortening. FEBS Lett, 453, 365–368.

17 von Zglinicki, T., Pilger, R. and Sitte, N. (2000) Accumulation of single-strand breaks is the major cause of telomere shortening in human fibroblasts. Free Radic Biol Med, 28, 64–74.

18 Starkweather, A.R., Alhaeeri, A.A., Montpetit, A., Brumelle, J., Filler, K., Montpetit, M., Mohanraj, L., Lyon, D.E. and Jackson-Cook, C.K. (2014) An integrative review of factors associated with telomere length and implications for biobehavioral research. Nurs Res, 63, 36–50.

19 Fouquerel, E., Parikh, D. and Opresko, P. (2016) DNA damage processing at telomeres: The ends justify the means. DNA Repair (Amst), 44, 159–168.

20 Barnes, R.P., Fouquerel, E. and Opresko, P.L. (2018) The impact of oxidative DNA damage and stress on telomere homeostasis. Mech Ageing Dev, in press.

21 Cadet, J., Wagner, J.R., Shafirovich, V. and Geacintov, N.E. (2014) One-electron oxidation reactions of purine and pyrimidine bases in cellular DNA. Int J Radiat Biol, 90, 423–432.

22 Nishimura, S. (2006) 8-Hydroxyguanine: From its discovery in 1983 to the present status. Proc Jpn Acad Ser B Phys Biol Sci, 82, 127–141.

23 Azzalin, C.M., Reichenbach, P., Khoriauli, L., Giulotto, E. and Lingner, J. (2007) Telomeric repeat containing RNA and RNA surveillance factors at mammalian chromosome ends. Science, 318, 798–801.

24 Chu, H.P., Cifuentes-Rojas, C., Kesner, B., Aeby, E., Lee, H.G., Wei, C., Oh, H.J., Boukhali, M., Haas, W. and Lee, J.T. (2017) TERRA RNA Antagonizes ATRX and Protects Telomeres. Cell, 170, 86–101.e116.

25 Arnoult, N., Van Beneden, A. and Decottignies, A. (2012) Telomere length regulates TERRA levels through increased trimethylation of telomeric H3K9 and HP1alpha. Nat Struct Mol Biol, 19, 948–956.

26 Graf, M., Bonetti, D., Lockhart, A., Serhal, K., Kellner, V., Maicher, A., Jolivet, P., Teixeira, M.T. and Luke, B. (2017) Telomere Length Determines TERRA and R-Loop Regulation through the Cell Cycle. Cell, 170, 72–85.e14.

27 James, S.J., Melnyk, S., Fuchs, G., Reid, T., Jernigan, S., Pavliv, O., Hubanks, A. and Gaylor, D.W. (2009) Efficacy of methylcobalamin and folinic acid treatment on glutathione redox status in children with autism. Am J Clin Nutr, 89, 425–430.

28 Chauhan, A. and Chauhan, V. (2006) Oxidative stress in autism. Pathophysiology, 13, 171–181.

29 James, S.J., Melnyk, S., Jernigan, S., Cleves, M.A., Halsted, C.H., Wong, D.H., Cutler, P., Bock, K., Boris, M., Bradstreet, J.J. et al. (2006) Metabolic endophenotype and related genotypes are associated with oxidative stress in children with autism. Am J Med Genet B Neuropsychiatr Genet, 141B, 947–956.

30 James, S.J., Rose, S., Melnyk, S., Jernigan, S., Blossom, S., Pavliv, O. and Gaylor, D.W. (2009) Cellular and mitochondrial glutathione redox imbalance in lymphoblastoid cells derived from children with autism. FASEB J, 23, 2374–2383.

31 Rose, S., Melnyk, S., Pavliv, O., Bai, S., Nick, T.G., Frye, R.E. and James, S.J. (2012) Evidence of oxidative damage and inflammation associated with low glutathione redox status in the autism brain. Transl Psychiatry, 2, e134.

32 Shpyleva, S., Ivanovsky, S., de Conti, A., Melnyk, S., Tryndyak, V., Beland, F.A., James, S.J. and Pogribny, I.P. (2014) Cerebellar oxidative DNA damage and altered DNA methylation in the BTBR T+tf/J mouse model of autism and similarities with human post mortem cerebellum. PLoS One, 9, e113712.

33 Panahi, Y., Salasar Moghaddam, F., Babaei, K., Eftekhar, M., Shervin Badv, R., Eskandari, M.R., Vafaee-Shahi, M., Pezeshk, H. and Pedram, M. (2020) Sexual Dimorphism in Telomere Length in Childhood Autism. *MedRxive*, doi: 10.1101/2020.04.30.20074765.

34 O’Callaghan, N., Baack, N., Sharif, R. and Fenech, M. (2011) A qPCR-based assay to quantify oxidized guanine and other FPG-sensitive base lesions within telomeric DNA. Biotechniques, 51, 403–411.

35 Panahi, Y., Salasar Moghaddam, F., Ghasemi, Z., Hadi Jafari, M., Shervin Badv, R., Eskandari, M.R. and Pedram, M. (2016) Selection of Suitable Reference Genes for Analysis of Salivary Transcriptome in Non-Syndromic Autistic Male Children. Int J Mol Sci, 17.

36 Sampl, S., Pramhas, S., Stern, C., Preusser, M., Marosi, C. and Holzmann, K. (2012) Expression of telomeres in astrocytoma WHO grade 2 to 4: TERRA level correlates with telomere length, telomerase activity, and advanced clinical grade. Transl Oncol, 5, 56–65.

37 Butler, M.G., Rafi, S.K. and Manzardo, A.M. (2015) High-resolution chromosome ideogram representation of currently recognized genes for autism spectrum disorders. Int J Mol Sci, 16, 6464–6495.

38 Kawanishi, S. and Oikawa, S. (2004) Mechanism of telomere shortening by oxidative stress. Ann N Y Acad Sci, 1019, 278–284.

39 Oikawa, S., Tada-Oikawa, S. and Kawanishi, S. (2001) Site-specific DNA damage at the GGG sequence by UVA involves acceleration of telomere shortening. Biochemistry, 40, 4763–4768.

40 Boiteux, S., O’Connor, T.R., Lederer, F., Gouyette, A. and Laval, J. (1990) Homogeneous Escherichia coli FPG protein. A DNA glycosylase which excises imidazole ring-opened purines and nicks DNA at apurinic/apyrimidinic sites. J Biol Chem, 265, 3916–3922.

41 Frenkel, K., Goldstein, M.S., Duker, N.J. and Teebor, G.W. (1981) Identification of the cis-thymine glycol moiety in oxidized deoxyribonucleic acid. Biochemistry, 20, 750–754.

42 Clark, J.M. and Beardsley, G.P. (1986) Thymine glycol lesions terminate chain elongation by DNA polymerase I in vitro. Nucleic Acids Res, 14, 737–749.

43 Vallabhaneni, H., O’Callaghan, N., Sidorova, J. and Liu, Y. (2013) Defective repair of oxidative base lesions by the DNA glycosylase Nth1 associates with multiple telomere defects. PLoS Genet, 9, e1003639.

44 Lee, H.T., Bose, A., Lee, C.Y., Opresko, P.L. and Myong, S. (2017) Molecular mechanisms by which oxidative DNA damage promotes telomerase activity. Nucleic Acids Res, 45, 11752–11765.

45 Luo, W., Muller, J.G., Rachlin, E.M. and Burrows, C.J. (2001) Characterization of hydantoin products from one-electron oxidation of 8-oxo-7,8-dihydroguanosine in a nucleoside model. Chem Res Toxicol, 14, 927–938.

46 Fleming, A.M., Ding, Y. and Burrows, C.J. (2017) Oxidative DNA damage is epigenetic by regulating gene transcription via base excision repair. Proc Natl Acad Sci U S A, 114, 2604–2609.

47 Park, J., Park, J.W., Oh, H., Maria, F.S., Kang, J. and Tian, X. (2016) Gene-Specific Assessment of Guanine Oxidation as an Epigenetic Modulator for Cardiac Specification of Mouse Embryonic Stem Cells. PLoS One, 11, e0155792.

48 Fouquerel, E., Lormand, J., Bose, A., Lee, H.T., Kim, G.S., Li, J., Sobol, R.W., Freudenthal, B.D., Myong, S. and Opresko, P.L. (2016) Oxidative guanine base damage regulates human telomerase activity. Nat Struct Mol Biol, 23, 1092–1100.

49 McNulty, J.M., Jerkovic, B., Bolton, P.H. and Basu, A.K. (1998) Replication inhibition and miscoding properties of DNA templates containing a site-specific cis-thymine glycol or urea residue. Chem Res Toxicol, 11, 666–673.

50 Bigler, E.D., Mortensen, S., Neeley, E.S., Ozonoff, S., Krasny, L., Johnson, M., Lu, J., Provencal, S.L., McMahon, W. and Lainhart, J.E. (2007) Superior temporal gyrus, language function, and autism. Dev Neuropsychol, 31, 217–238.

51 Karikkineth, A.C., Scheibye-Knudsen, M., Fivenson, E., Croteau, D.L. and Bohr, V.A. (2017) Cockayne syndrome: Clinical features, model systems and pathways. Ageing Res Rev, 33, 3–17.

52 Batenburg, N.L., Mitchell, T.R., Leach, D.M., Rainbow, A.J. and Zhu, X.D. (2012) Cockayne Syndrome group B protein interacts with TRF2 and regulates telomere length and stability. Nucleic Acids Res, 40, 9661–9674.

53 Sagie, S., Edni, O., Weinberg, J., Toubiana, S., Kozlovski, T., Frostig, T., Katzin, N., Bar-Am, I. and Selig, S. (2017) Non-random length distribution of individual telomeres in immunodeficiency, centromeric instability and facial anomalies syndrome, type I. Human molecular genetics, 26, 4244–4256.

54 Jung, H., Park, H., Choi, Y., Kang, H., Lee, E., Kweon, H., Roh, J.D., Ellegood, J., Choi, W., Kang, J. et al. (2018) Sexually dimorphic behavior, neuronal activity, and gene expression in Chd8-mutant mice. Nat Neurosci, 21, 1218–1228.

55 Lord, C., Pickles, A., McLennan, J., Rutter, M., Bregman, J., Folstein, S., Fombonne, E., Leboyer, M. and Minshew, N. (1997) Diagnosing autism: analyses of data from the Autism Diagnostic Interview. J Autism Dev Disord, 27, 501–517.

56 Krokan, H.E. and Bjoras, M. (2013) Base excision repair. Cold Spring Harb Perspect Biol, 5, a012583.

57 Cawthon, R.M. (2002) Telomere measurement by quantitative PCR. Nucleic Acids Res, 30, e47.

58 Karakaya, A., Jaruga, P., Bohr, V.A., Grollman, A.P. and Dizdaroglu, M. (1997) Kinetics of excision of purine lesions from DNA by Escherichia coli Fpg protein. Nucleic Acids Res, 25, 474–479.

59 O’Callaghan, N., Dhillon, V., Thomas, P. and Fenech, M. (2008) A quantitative real-time PCR method for absolute telomere length. Biotechniques, 44, 807–809.

60 Cawthon, R.M. (2009) Telomere length measurement by a novel monochrome multiplex quantitative PCR method. Nucleic Acids Res, 37, e21.

61 Deng, Z., Wang, Z., Stong, N., Plasschaert, R., Moczan, A., Chen, H.S., Hu, S., Wikramasinghe, P., Davuluri, R.V., Bartolomei, M.S. et al. (2012) A role for CTCF and cohesin in subtelomere chromatin organization, TERRA transcription, and telomere end protection. EMBO J, 31, 4165–4178.

